# A population-based cohort study to establish clinical characteristics and outcomes of patients experiencing methicillin-sensitive *Staphylococcus aureus* bacteremia as a function of the cefazolin high inoculum effect

**DOI:** 10.64898/2025.12.03.25341560

**Authors:** Alexander J. Kipp, Kristine Du, Barbara Wadell, Stephen Robinson, Julianna Svishchuk, John M. Conly, Annegret Ulke-Lemee, Maryam Mapar, Ian Lewis, Daniel B. Gregson, Michael D. Parkins

**Affiliations:** The University of Calgary, Department of Pathology and Laboratory Medicine, Calgary, Alberta, Canada; The University of Calgary, Department of Microbiology, Immunology & Infectious Diseases, Calgary, Alberta, Canada; The University of Calgary, Department of Medicine, Calgary, Alberta, Canada; Dalhousie University, Department of Medicine, St John, New Brunswick, Canada; The University of Calgary, Department of Biological Sciences, Calgary, Alberta, Canada

**Keywords:** *S. aureus*, *Staphylococcus aureus*, high-inoculum effect, cefazolin, cloxacilli*n*, *blaZ*, penicillinase, clonal complex, bacteremia

## Abstract

**Background:** *Staphylococcus aureus* bacteremia is a leading cause of morbidity and mortality. Several phenotypes (e.g. methicillin-resistance) influence patient outcomes. The high inoculum effect (HIE) is characterized by reduced susceptibility to beta-lactam antibiotics, most notably cefazolin, at high inoculums *in-vitro*.

**Methods:** A population-based cohort was used to assess all MSSA bacteremia in Calgary, Alberta, from 2012-14 and 2019 (n=1.5 million). Isolates underwent genomic sequencing and cefazolin susceptibility testing at 10^5^ and 10^7^ CFU/ml where the HIE was defined as a 4X-increase in minimum-inhibitory concentration (MIC) and pronounced (PIE) defined as MIC≥16 ug/ml at 10^7^ CFU/ml.

**Results:** The incidence of MSSA bacteremia with HIE decreased from 38.9 to 24.2% between the two time periods. Patients infected with HIE phenotype could not be differentiated based on demographics, source of bacteremia, or clinical biomarkers at presentation. Sequencing confirmed associations of HIE with *blaZ* A, *agr3,* and clonal complex 30. HIE was not associated with outcomes including clearance-time and all-cause mortality when assessed in aggregate or as a function of treatment. Relapses, however, were only documented with cefazolin. PIE was observed in 3.7% of isolates and was associated with significant increased all-cause 180-day mortality, irrespective of treatment, but not at one year.

**Discussion:** The HIE, but not PIE, is common in an unselected general population cohort. Neither demographics nor clinical biomarkers can be used to predict HIE. If there is a deleterious impact of this phenotype on patient outcomes, it is modest and may be masked by empiric therapy provided prior to MSSA bacteremia confirmation.

**Importance Statement:** A prospective cohort study by Miller *et al.* (2018) observed a significant increase in 30-day mortality in individuals experiencing methicillin-sensitive *Staphylococcus aureus* (MSSA) bacteremia, with isolates exhibiting the high-inoculum effect (HIE) phenotype, when treated with the cefazolin. Our study sought to understand the epidemiology and impact of the HIE, using a population-based study design thereby mitigating the selection bias associated with other conventional cohort studies (focused on specific hospitals, at-risk groups, or clinics). We address the effects of HIE on clinical outcomes, predictive factors, and associated genomic contributors for the HIE phenotype within an unbiased population. These data are important for clinicians by highlighting if it is possible to predict HIE phenotype based on clinical and genomic factors, and to establish if cefazolin use associates with worse outcomes when MSSA causing bacteremia exhibits the HIE phenotype.

## Introduction

*Staphylococcus aureus* blood stream infections (BSI) are a significant cause of morbidity and mortality. Several *S. aureus* phenotypes influence patient outcomes, including methicillin resistance (MRSA), small-colony variants (SCVs), and to a lesser degree Panton-Valentine leucocidin toxin (PVL) (1–3). One phenotype increasingly explored is the high inoculum effect (HIE) to beta-lactam antibiotics. HIE was first discovered through testing of cephaloridine and cephalothin in the 1960s (4,5). When specific *S. aureus* strains were present at high inoculum, there was reduced killing at 18hrs compared to methicillin and quinacillin (4,5). Much research has since focused on cefazolin owing to its susceptibility to this phenotype and increased use as first-line therapy. The HIE has primarily been attributed to the *S. aureus* beta-lactamase gene *blaZ* types A and C, likely mediated through partial hydrolysis of beta-lactams (6–14) and polymorphisms in the accessory gene regulating region *agr* known a*s agr3* (13–16). These alleles are also associated with the *S. aureus* clonal complex 30 (CC), a lineage associated with persistent infections and metastatic complications (8,9,17–19) in methicillin-sensitive *S. aureus* (MSSA) and methicillin-resistant (MRSA) (20,21).

With potential inferiority of cefazolin in the context of HIE, and supporting *in-vivo* animal studies (22,23), there exists debate about best management of MSSA BSI despite cefazolin’s preferable safety and tolerability profile (24). Studies have linked HIE with increased risk of deep-seated infections and mortality (7,14,23,25). A recent study by Miller *et al*. (2018) showed that patients treated with cefazolin as first-line therapy had higher 30-day mortality when the *S. aureus* strain exhibited the HIE phenotype. Others groups, however, were unable to link HIE to increased mortality (9,26,27).

We sought to establish the prevalence, risk factors, and outcomes of MSSA BSI in a longitudinal, non-selected cohort of 1.5 million individuals in a high-income country, as a function of HIE phenotype. As a secondary objective, a subset of patients had their MSSA isolates genotyped which were used to assess for associations of the HIE phenotype to *blaZ*, *agr* alleles, and CC.

A population-based study design was used as it is most appropriate to understand the epidemiology and outcomes of the HIE (28). By capturing all episodes of disease occurring within a well-defined adult population, we aimed to reduce selection bias. Such a cohort study assessing HIE has not previously been performed.

## Methods

### Patients and Clinical Epidemiology

All patients experiencing MSSA BSIs in Calgary were identified over the years 2012-2014 and 2019, using the Alberta Provincial Laboratories laboratory information system which captures all lab-work performed in Alberta, Canada. Cohort inclusion criteria included age ≥18 years and residence within the 2014 Calgary Health Zone based on primary residence postal code. Only first episodes of bacteremia were included in any year, and relapses and recurrences were not counted separately when calculating incidence. Patients were followed to 1-year post-bacteremia for statistical and chart review purposes. Patients who immediately left against medical advice or had no incident clinical assessment recorded were excluded. Methodology was adapted from Lam *et al.* (2019). Ethics approval was obtained from the Conjoint Regional Health Ethics Board (REB14-1456).

The Calgary Health Zone encompasses the metropolitan area of Calgary and its surrounding communities (Airdrie, Cochrane, and Okotoks), including a total population of 1.5 million in 2019. All acute care within is provided by Alberta Health Services (AHS), the provincial health authority. Geographically this includes four adult hospitals with over 2500 dedicated inpatient beds (29). Linked patient admission demographics, epidemiological characteristics, treatments, and disease outcomes were extracted electronically and supplemented with a chart audit. General population data for the catchment area were used to determine yearly incident rates of MSSA BSI (30). The civic census was collected every 5 years (e.g. 2014, 2019) and does not have an age breakdown delineating those ≥18 years of age, therefore inferences were made from the 2014 data as previously outlined (31) to consider only 82% of the population as ≥18.

### Laboratory Testing

Blood cultures were performed using the BacTAlert^®^ Virtuo^®^ (bioMerieux Canada, St. Laurent, Quebec) automated blood culture system. Isolates were identified as *S. aureus* through minimal biochemical characteristics identification as outlined in CLSI M35 (2008), along with Vitek 2^®^ ID and Vitek^®^ MS (bioMerieux Canada, St. Laurent, Quebec) in later years. Isolates were determined methicillin-sensitive utilizing Vitek^®^ 2 AST (bioMerieux Canada, St. Laurent, Quebec) testing with screening provisions for oxacillin and cefoxitin according to CLSI M100 Ed. 29 (2019).

836 unique MSSA isolates underwent testing for the HIE phenotype utilizing broth microdilution following methods adapted from Nanni *et al.* (2003), Livorsi *et al.* (2012), and Wang et al. (2018). Isolates were defined as HIE+ if they had a ≥4-fold difference in minimum-inhibitory concentration (MIC) at high-inoculum (HI: 5x10^7^ CFU/mL) relative to standard inoculum (SI: 5x10^5^ CFU/mL). A secondary outcome was the occurrence of pronounced inoculum effect (PIE) defined as when the cefazolin MIC at 5x10^7^ CFU/mL was ≥16ug/mL, a since revised CLSI breakpoint defining resistance in *S. aureus* (7,14,23,27).

MSSA BSIs were categorized as community acquired (CA), healthcare associated (HCA), or hospital acquired (HA) using definitions from Friedman *et al.* (2002) and Lenz *et al.* (2012). MSSA BSI was further subcategorized as “complicated” or “uncomplicated” based on definitions from IDSA and CMAJ (32,33). Sources of infection and metastatic complications were identified from sterile fluids when applicable (i.e., joint aspirates) and utilizing clinical summaries as described previously (31). Infectious endocarditis was defined using the modified Duke’s Criteria (32) with both *definite* and *probable* being included as positive cases for statistical purposes. Ruling out infective endocarditis as part of the definition of “uncomplicated bacteremia” necessitated a trans-esophageal echocardiogram. Hematologic and biochemical results were evaluated based on those closest in proximity to the incident positive blood culture.

Recurrences were defined as a repeat MSSA BSI after documenting clearance via blood cultures or as identified by the most-responsible provider or infectious disease physician overseeing the case, and completion of therapy according to IDSA guidelines. They were further categorized as relapses occurring in <90 days, <180 days, and <1 year. When documenting blood culture clearance at 24hrs, 48hrs, and 72hrs these were non-inclusive and required no other positive blood cultures for MSSA BSI within the current illness.

A subset of isolates (n=599, 71.7%) underwent whole genome sequencing to determine their *blaZ* type, *agr* allele, and CC. Bacterial DNA was purified after lysis with Lysozyme L6876, Lysostaphin L7386, Proteinase K 70663 and RNAse A R5503 (Sigma-Aldrich Canada, Oakville, Ontario) using the MagMax DNA purification kit AM1840 (Thermo Fisher Scientific, Waltham, MA). DNA was quantified using the DNA Quant-it dsDNA assay kit Q33120 (Thermo Fisher Scientific, Waltham, MA). Sequencing libraries were generated using the Nextera XT DNA library prep kit (Illumina, San Diego, CA) and 151x151 bp paired-end sequencing was performed on the Illumina HiSeqX platform.

### Bioinformatic analysis

Assembly of genomes and identification of antimicrobial resistance genes was performed on computing infrastructure at the University of Calgary using open-source computational pipeline seQuoia (34) – leveraging the Calgary BSI Cohort – and are accessible at http://www.resistancedb.org/. The median sequencing depth of isolates was 165X. Briefly, adaptor sequences were trimmed using TrimGalore (35). Strain Genome Search Tool StrainGST (36) was used to find closest reference genomes and alignment was done using Burrows-Wheeler Aligner (37). Variant calling files were generated using Pilon (38). Isolates with assembled sequences missing *blaZ* but positive for HIE were checked by *de novo* assembly SKESA assembler (39) and confirmed with breseq SNP calling. Genomic data and that from a clinical chart audit were merged using Microsoft Excel PowerQuery (Redmond, WA). Entries were matched using study specific patient identification number (PID). Duplicates were formed by the software when a PID had multiple independent blood cultures, drawn the same day or had a recurrent infection. Duplicate PIDs were assessed with a chart audit to confirm validity based on blood culture collection time.

### Statistical analysis

Analyses were performed with Graph Pad Prism 10.4.1 (Boston, MA). Continuous variables were presented as medians with interquartile ranges. Non-normally distributed data were then evaluated using Mann-Whitney U test. Binary variables were first compared using a Fischer’s exact test. Risk ratios were calculated by dividing the proportion with the independent variable by the proportion without it and reported using 95% confidence intervals (CI). A logistic regression model identified independent factors associated with the HIE phenotype. Factors identified through univariate analysis (p ≤0.10) were initially included and then subject to manual backward stepwise variable elimination to develop an efficient model. 1-year survival was calculated using Mantel-Cox Kaplan-Meier curves with all-cause mortality or composite all-cause mortality and relapse. Results are reported as odds ratios (OR), relative risk (RR) or hazard ratio (HR) with 95% CI. Significance was determined by a p-value ≤0.05.

## Results

### Incidence and Epidemiology of MSSA Bacteremia and HIE in general populations

There were 176, 229, 251, and 215 cases of MSSA bacteremia within the Calgary health region in 2012, 2013, 2014, and 2019, respectively (total=871). The population adjusted rate of MSSA bacteremia from the 2012-2014 cohort as previously reported (31) decreased in the 2019 cohort (Figure 1) from 25.6 in 2014 to 20.6 in 2019 (p=0.02) and was on average 22.4 cases per 100,000 residents. Of the 871 total isolates included, 96% (n=836) were recoverable from Alberta Precision Laboratory Biobank and subjected to HIE testing. BSI Isolates that failed to be recovered (n=33) did not differ in respect to patient characteristics including median age or sex (p>0.05) and the proportion of recovered isolates was above 93% in every study year with a maximum of 100% in 2019.

**Figure 1:**
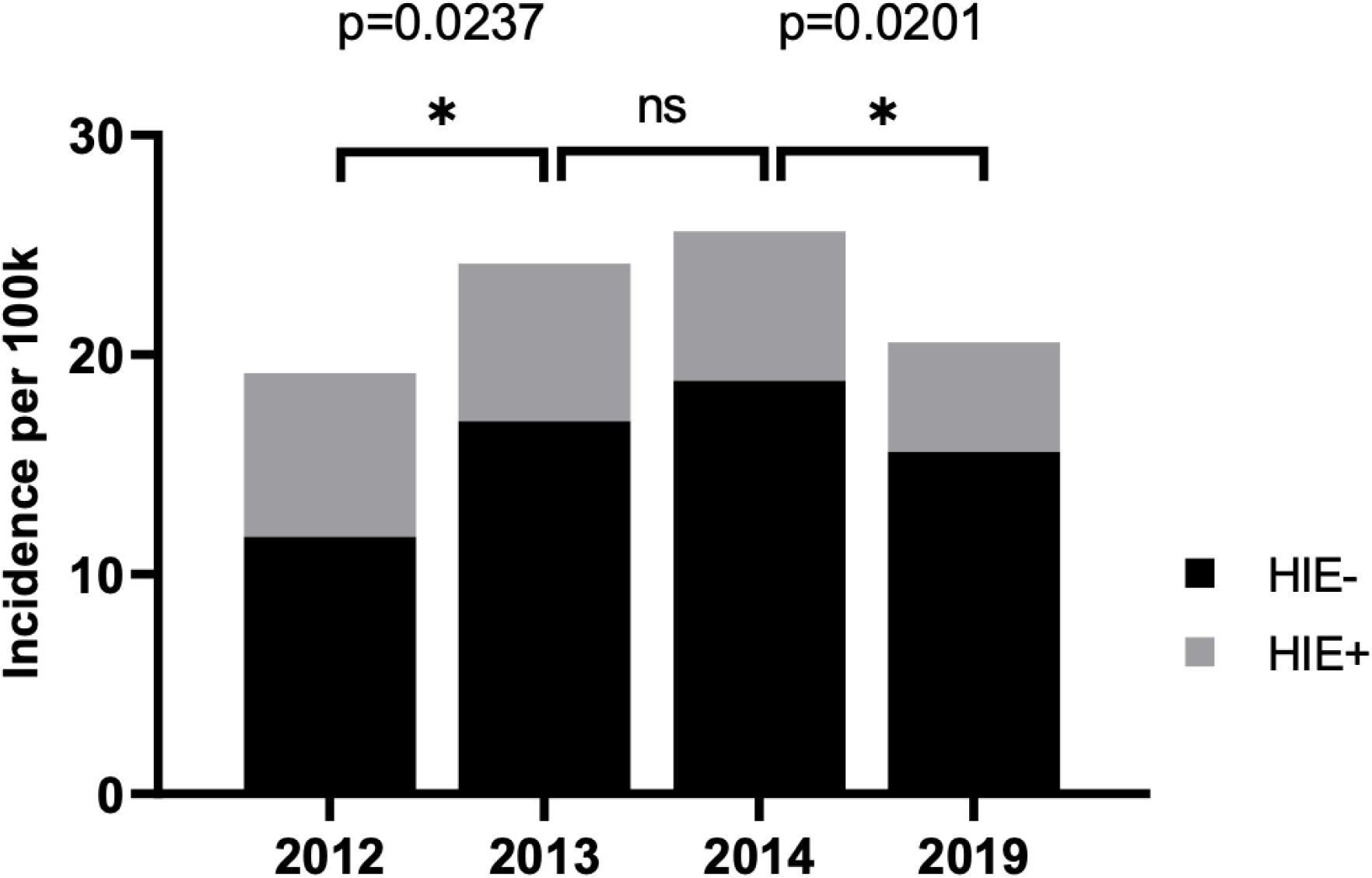
Incidence of MSSA BSI as a function of cefazolin high inoculum effect (HIE) status.

The proportion of MSSA BSI isolates demonstrating HIE differed over the study periods showing an overall significant decrease in incidence from 38.9% in 2012 to 24.4% in 2019 (p=0.003) but not individual inter-years (p>0.05). This corresponded to an extrapolated average HIE BSI incidence of 6.6/100,000 residents (SD = 1.12) assuming the fraction of HIE+ isolates were representative of the population. In contrast, the proportion of isolates exhibiting PIE was markedly lower at 30/836 (3.59%), corresponding to an incidence rate of 0.79/100,000 residents. All isolates that had PIE also displayed HIE.

Cases of MSSA BSI were predominately community acquired (55%) versus healthcare-associated (20%) and nosocomial (25%). This proportion did not differ significantly from year-to-year (p=0.46). Mode of acquisition was not associated with HIE (Table 1).

**Table 1:**
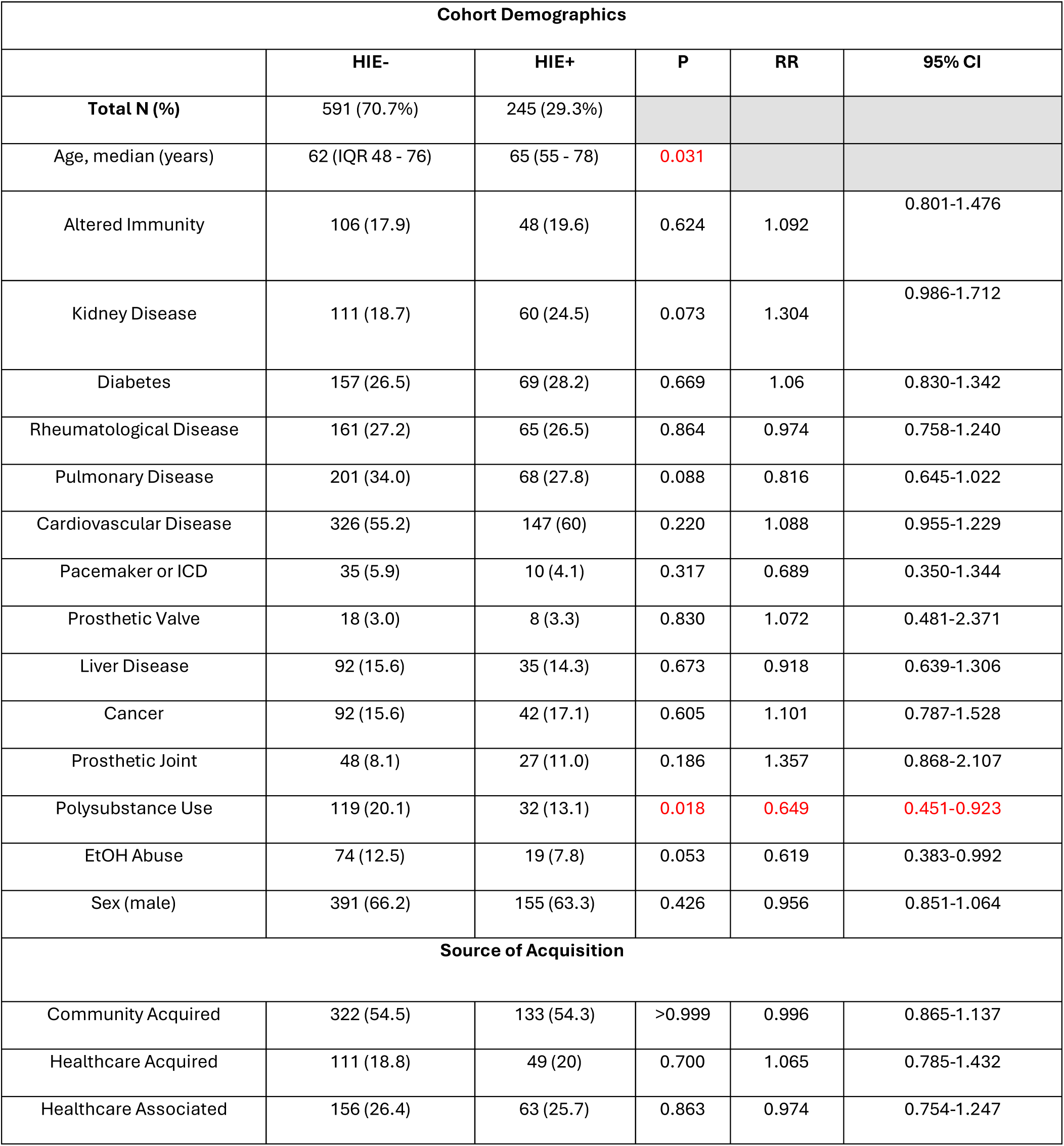
Clinical characteristics of those with MSSA BSI as a function of HIE.

| Cohort Demographics |  |  |  |  |  |
| --- | --- | --- | --- | --- | --- |
|  | HIE- | HIE+ | P | RR | 95% CI |
| **Total N (%)** | 591 (70.7%) | 245 (29.3%) |  |  |  |
| Age, median (years) | 62 (IQR 48 - 76) | 65 (55 - 78) | 0.031 |  |  |
| Altered Immunity | 106 (17.9) | 48 (19.6) | 0.624 | 1.092 | 0.801-1.476 |
| Kidney Disease | 111 (18.7) | 60 (24.5) | 0.073 | 1.304 | 0.986-1.712 |
| Diabetes | 157 (26.5) | 69 (28.2) | 0.669 | 1.06 | 0.830-1.342 |
| Rheumatological Disease | 161 (27.2) | 65 (26.5) | 0.864 | 0.974 | 0.758-1.240 |
| Pulmonary Disease | 201 (34.0) | 68 (27.8) | 0.088 | 0.816 | 0.645-1.022 |
| Cardiovascular Disease | 326 (55.2) | 147 (60) | 0.220 | 1.088 | 0.955-1.229 |
| Pacemaker or ICD | 35 (5.9) | 10 (4.1) | 0.317 | 0.689 | 0.350-1.344 |
| Prosthetic Valve | 18 (3.0) | 8 (3.3) | 0.830 | 1.072 | 0.481-2.371 |
| Liver Disease | 92 (15.6) | 35 (14.3) | 0.673 | 0.918 | 0.639-1.306 |
| Cancer | 92 (15.6) | 42 (17.1) | 0.605 | 1.101 | 0.787-1.528 |
| Prosthetic Joint | 48 (8.1) | 27 (11.0) | 0.186 | 1.357 | 0.868-2.107 |
| Polysubstance Use | 119 (20.1) | 32 (13.1) | 0.018 | 0.649 | 0.451-0.923 |
| EtOH Abuse | 74 (12.5) | 19 (7.8) | 0.053 | 0.619 | 0.383-0.992 |
| Sex (male) | 391 (66.2) | 155 (63.3) | 0.426 | 0.956 | 0.851-1.064 |
| Source of Acquisition |  |  |  |  |  |
| Community Acquired | 322 (54.5) | 133 (54.3) | >0.999 | 0.996 | 0.865-1.137 |
| Healthcare Acquired | 111 (18.8) | 49 (20) | 0.700 | 1.065 | 0.785-1.432 |
| Healthcare Associated | 156 (26.4) | 63 (25.7) | 0.863 | 0.974 | 0.754-1.247 |

The most common comorbidity in the cohort was cardiovascular disease (57%), then pulmonary disease (32%) and diabetes (29%). The cohort was predominantly male (65%). Patient characteristics generally did not differ amongst individuals experiencing bacteremia with HIE+ MSSA except for median age and substance use (Table 1), whereby risk of HIE+ MSSA BSI increased with age and decreased with polysubstance use history.

### Clinical Markers and their Association to HIE

The common sources of MSSA BSI were determined to be unknown (39.9%), skin and soft tissue infection (21.5%), and pneumonia (11.7%). The sources of bacteremia only differ based on HIE status for septic arthritis, where HIE+ was observed more commonly (Table 2A).

**Table 2:** Characteristics of MSSA BSI at initial presentation as a function of HIE **(A).** Sources of MSSA bacteremia. **(B).** Biochemical markers of inflammation.

| Source of Bacteremia and HIE (A) |  |  |  |  |  |
| --- | --- | --- | --- | --- | --- |
|  | HIE- | HIE+ | P | RR | 95% CI |
| **Total N (%)** | 591 (70.7%) | 245 (29.3%) |  |  |  |
| Unknown | 237 (40.1) | 99 (40.4) | 0.938 | 1.014 | 0.843-1.208 |
| SSTI | 137 (23.2) | 50 (20.4) | 0.310 | 0.852 | 0.631-1.141 |
| Pneumonia | 68 (11.5) | 25 (10.2) | 0.907 | 0.965 | 0.638-1.446 |
| Peritonitis | 4 (0.7) | 0 | >0.999 | 1.206 | 0.259-5.584 |
| PVC | 10 (1.7) | 4 (1.6) | 0.766 | 0.724 | 0.216-2.407 |
| CVC | 43 (7.3) | 17 (6.9) | 0.884 | 0.932 | 0.545-1.581 |
| Surgical | 10 (1.7) | 6 (2.4) | 0.432 | 1.535 | 0.620-3.782 |
| Septic Arthritis | 27 (4.6) | 22 (9.0) | 0.014 | 2.041 | 1.185-3.499 |
| Urine | 11 (1.9) | 2 (0.8) | 0.306 | 0.452 | 0.142-1.430 |
| Pacemaker | 4 (0.7) | 2 (0.8) | >0.999 | 0.689 | 0.163-2.887 |
| Other | 19 (3.2) | 6 (2.4) | >0.999 | 1.072 | 0.352-3.241 |
| Intravascular | 3 (0.5) | 0 | 0.327 | 0 | 0.000-2.298 |
| Osteomyelitis | 15 (2.5) | 9 (3.7) | 0.369 | 1.447 | 0.654-3.185 |
| Markers of HIE (B) |  |  |  |  |  |
| WBC Count, median | 12.1 (8.3 - 17.3) | 12.65 (8.3 - 17.35) | 0.788 |  |  |
| PLT, median | 196 (122 - 274.3) | 193 (122 - 303) | 0.523 |  |  |
| CRP, median | 162.5<br>(82.75 - 271.7) | 141.5<br>(50.73 - 269.9) | 0.861 |  |  |
| Bandemia | 78 (13.2) | 32 (13.1) | >0.999 | 0.9896 | 0.673-1.442 |
| Toxic Changes | 103 (17.4) | 34 (13.9) | 0.220 | 0.7963 | 0.555-1.131 |

Laboratory biomarkers drawn at presentation to assess inflammatory response (e.g., WBC, CRP) did not differ in patients with HIE+ MSSA compared to those without (Table 2B). When metrics that might reasonably serve as a biomarker to predict risk of HIE were assessed by multivariable regression, septic arthritis showed a significant association with HIE+ (Figure 2); however, the overall proportion of cases linked to septic arthritis were low (5.9%). Similar analyses were conducted for PIE isolates, and no significant differences were observed (p>0.05, data not shown).

**Figure 2:**
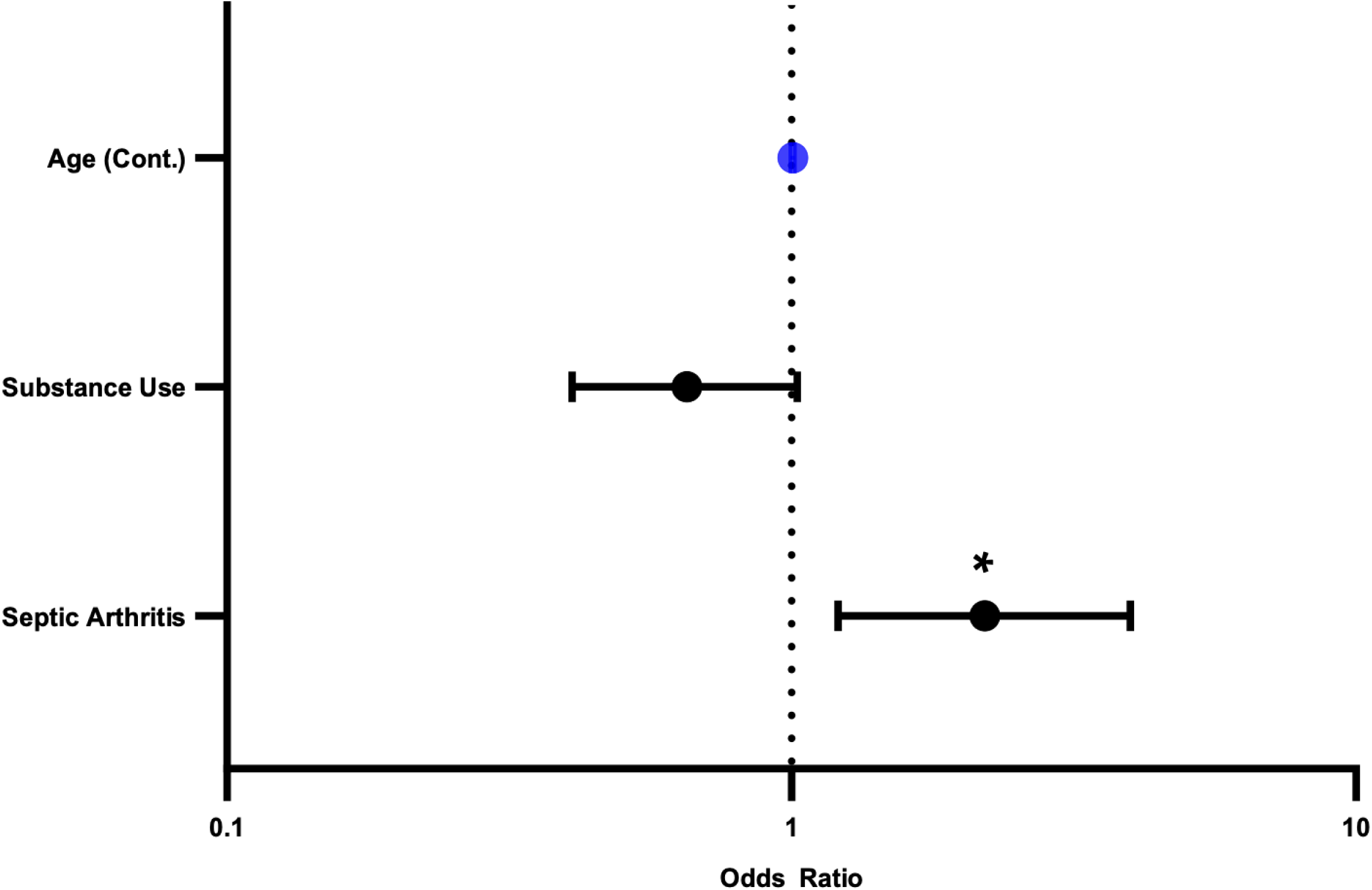
Clinical characteristics that associate with MSSA BSI as a function of the HIE through multivariate analysis. Blue represents a continuous variable.

### Influence of HIE on patient outcomes irrespective of treatments received

There was a total of 177 cohort deaths by 30 days. There were no differences in 30-day mortality based on HIE status (HIE+ 21.2% vs HIE- 20.3%, p=0.778). There was a greater increase in mortality at 180-days of 34.7% for HIE+ vs 28.3% HIE-, however this did not reach significance (p=0.069). There was no overall survival difference measured at one year (HIE+ 60.8% vs HIE- 67.9%, p=0.076, Figure 3) or as a composite measurement of all-cause mortality and relapses (p=0.09, Figure 4). There was no association between HIE status and the need for ICU support (p=0.194).

**Figure 3:**
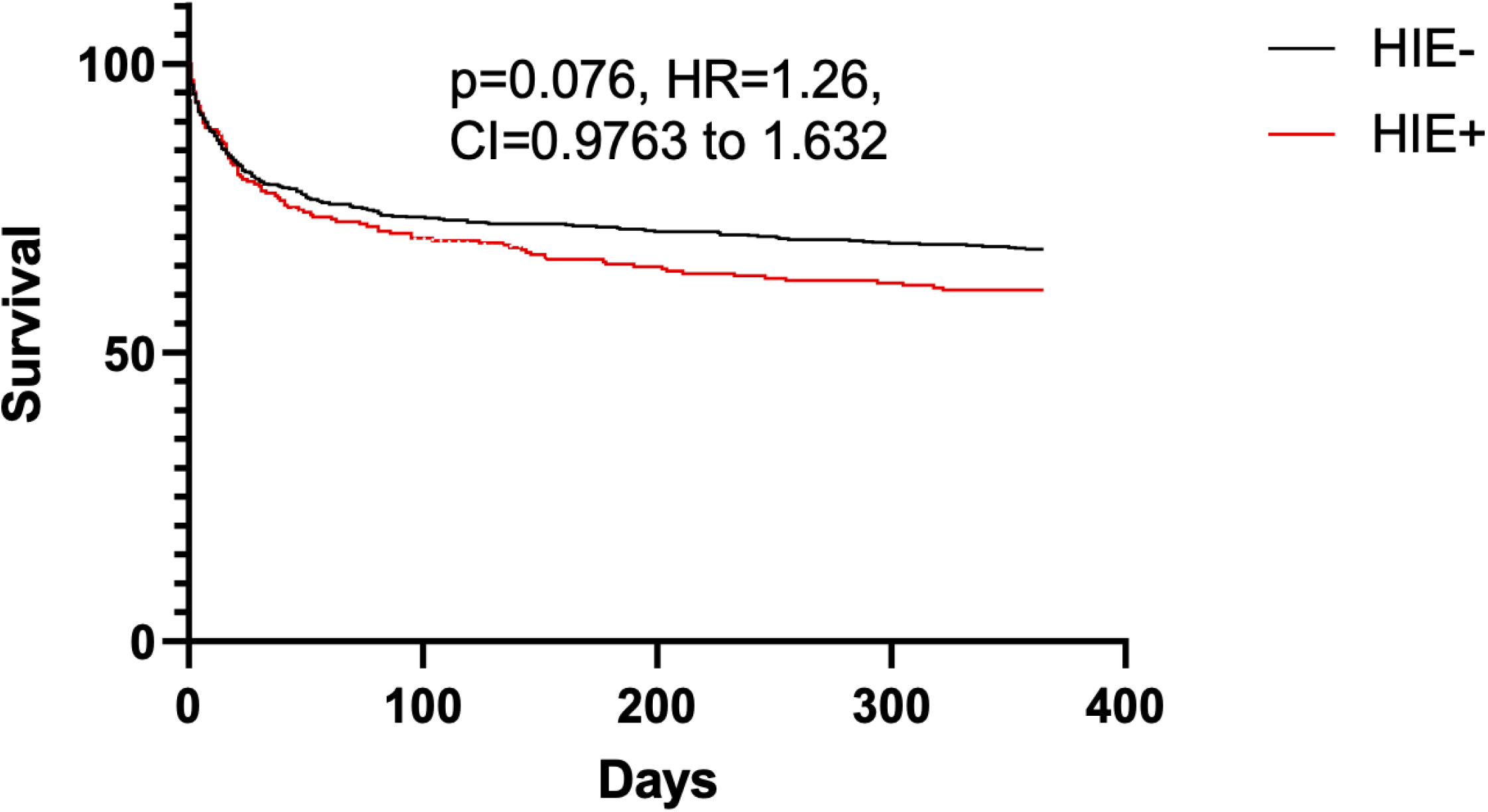
One year survival curve for those bacteremic with MSSA with the HIE (red) and without (black).

**Figure 4:**
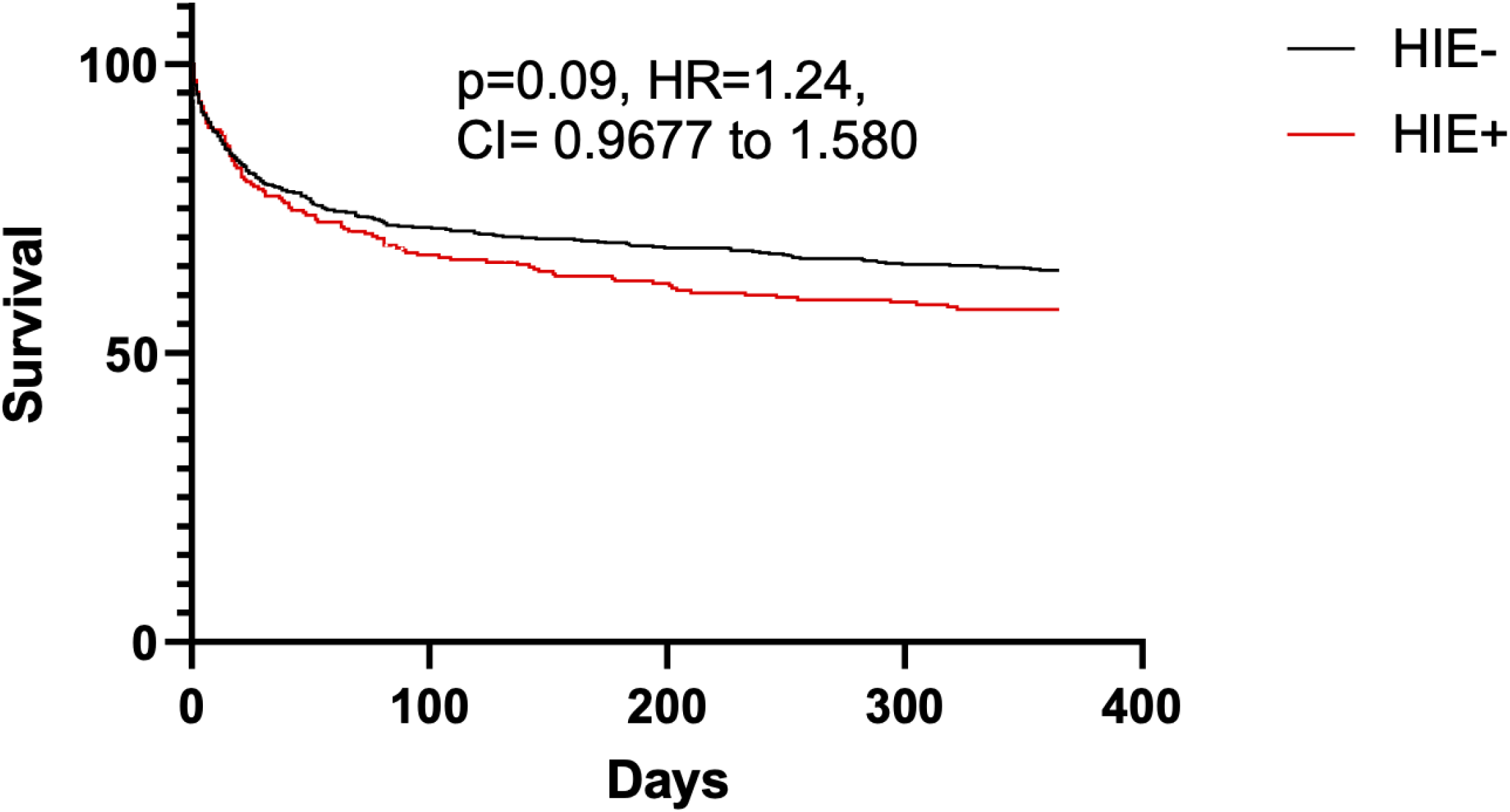
One year composite outcome curve for those infected with MSSA with HIE (red) and without (black) including all-cause mortality and relapse.

Most of the cohort classified as complicated MSSA BSI (84%) and under half (44%) had probable or definite evidence of infective endocarditis, of which 70.7% received TTE and 22.1% trans-esophageal echocardiogram (TEE). The presence of isolates expressing HIE did not associate with either outcome (Table 3). Only 11% of the cohort was identified as having a metastatic complication of MSSA BSI and did not differ by HIE status (p=0.336).

**Table 3:**
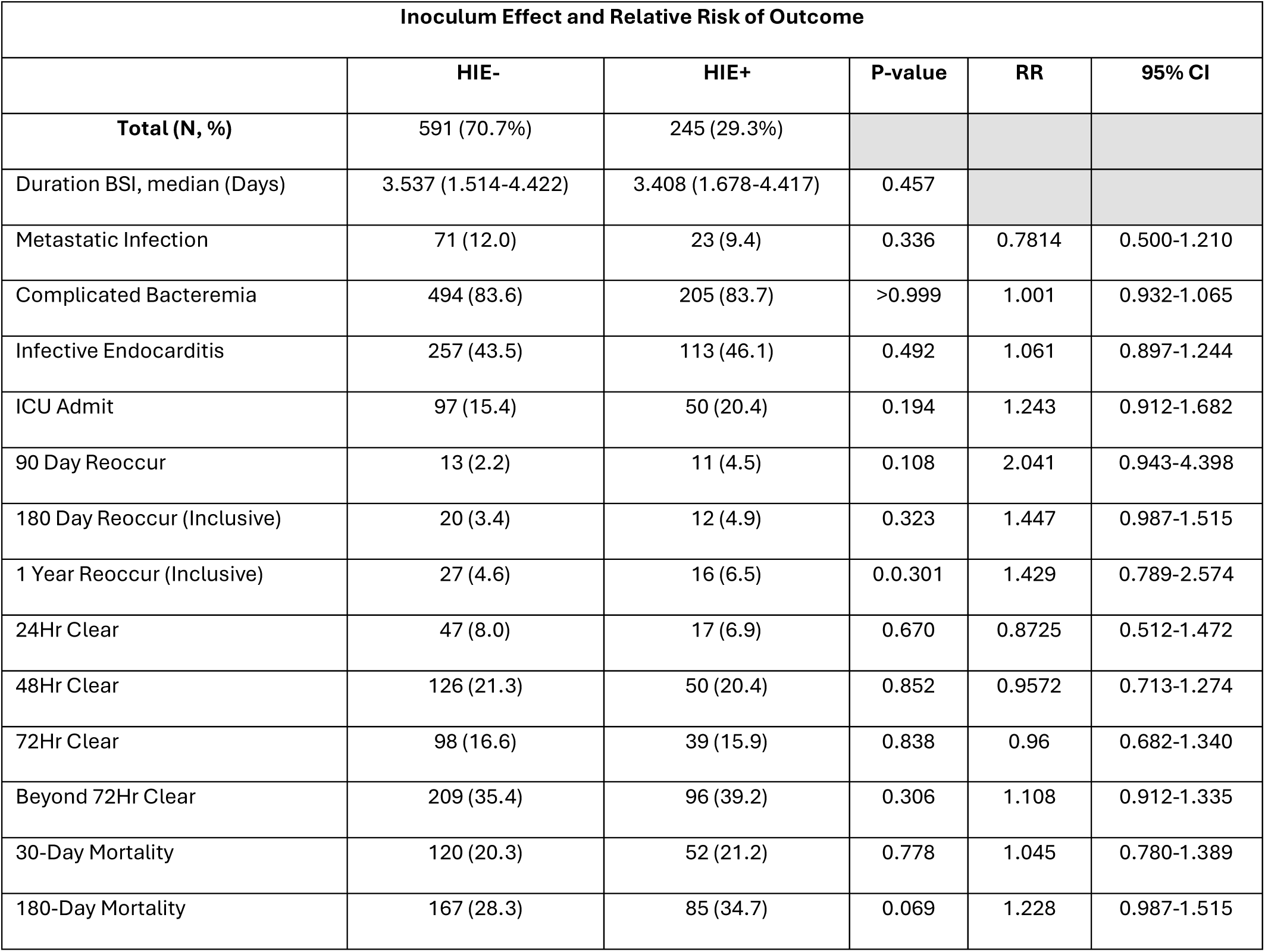
Outcomes of MSSA BSI as a function of HIE status.

Total MSSA BSI recurrence rates were low (2.8% at 90-days, 3.8% at 180-days, 5.1% at 1-year). Recurrence rates were not influenced by HIE status (p>0.05). Duration of MSSA BSI did not differ between patients based on HIE status, with a median of 3.54 days (IQR 1.533-4.36) for HIE+ vs 3.41 days (IQR 1.68-4.42) for HIE- (p=0.457). Similarly, the likelihood of clearance of BSI by 24hrs, 48hrs, and 72hrs was not influenced by HIE status (Table 3).

Similar analyses were conducted for PIE isolates. The notable difference observed was an increase in 180-day mortality in those with PIE (HIE+ 50% vs HIE- 29.4%, p=0.024, RR=1.7) however, there was no survival difference at 1-year (p=0.087, Supplemental Figure 1).

### Influence of HIE on patient outcomes as a function of treatments received

Chart audit of treatments showed remarkable variability. Within the first 24hrs of identification of positive blood cultures, most patients received two antimicrobial classes (74.2%), 24.5% three classes, and 6.8% four or more classes. Specifically, 72.5% of patients received vancomycin, whereas potent anti-Staphylococcal beta-lactam therapy (defined as cefazolin, carbapenem, cloxacillin but not ceftriaxone or piperacillin-tazobactam) were administered to 23%.

For patients who had either cefazolin or cloxacillin as their definitive agent at 48hrs, a significant 30-day mortality was favored in cefazolin (Cefazolin 11.2% vs Cloxacillin 24.4%, p=0.041, RR=0.459). This did not remain significant at/beyond 180-day nor was there a survival difference at 1-year (Supplemental Figure 2). There was a trend towards increased risk of those with metastatic complications being treated with cloxacillin, however, this did not reach significance (Cefazolin 8.7% vs Cloxacillin 17%, p=0.149, Supplemental Table 2).

Of patients with HIE+ MSSA BSI who received ≥4 weeks of total treatment and at least one parenteral anti-Staphylococcal beta-lactam was used for ≥3 weeks - specifically, cefazolin (75.2%, n=79) or cloxacillin (24.7%, n=26). Treatment duration did not differ based on regimen (median, cefazolin 44.10 days IQR 34.86-51.10, cloxacillin 44.72 days IQR 32.31-53.11, p=0.675). Mortality at 180-days did not differ based on receipt of either therapy (p=0.376). Recurrence rates at 90- and 180-days did not differ, however, only patients receiving cefazolin had recurrence by 1-year (11.4% vs 0%, p=0.196, Supplemental Table 2). A higher rate of treatment discontinuation secondary to adverse events (e.g., allergic reactions, side effects) requiring rotation to alternate antimicrobial regimens occurred in cloxacillin treatments compared to cefazolin (19.3% vs 11.3%, p=0.017, Supplemental Table 3). A 1-year survival curve was not significant in HIE+ when controlling treatment and duration (HR=1.09, p=0.852, Supplemental Figure 3).

### Genotypic Characteristics of MSSA Isolates with HIE Phenotype

Of 836 isolates that had HIE testing, a total of 599 underwent WGS to characterize *blaZ*, *agr* genes, and MSSA CC. This included 131 (78.4%) in 2012, 198 (89.2%) in 2013, 204 (87.2%) in 2014, and 66 (30.7%) in 2019. BSI Isolates that underwent WGS did not differ in patient characteristics including median age or sex (p>0.05) from those that were not assessed.

30.4% of isolates were identified as *blaZ* A, 12.7% B, 24% C, 0.3% D, and 32.6% were unknown. Incidences differed by study period, with a decrease of 27% of *blaZ* A from 2012 to 2019 and a corresponding increase of 11.9% for *blaZ* C, 13.4% *blaZ* B, and 18.8% unknown. Presence of *blaZ* A significantly associated with HIE+ (p=<0.001, RR=3.88) and PIE+ (p=<0.001, RR=2.90), whereas *blaZ* B and unknown types were negatively associated with HIE+ but not PIE+ (Table 4).

**Table 4:** HIE and PIE univariate association to *blaZ* and *agr* alleles.

| Univariate Association to *blaZ*, *agr* in HIE |  |  |  |  |  |
| --- | --- | --- | --- | --- | --- |
|  | HIE- | HIE+ | P-Value | RR | 95% CI |
| **Total N (%)** | 427 (71.3) | 172 (28.7) |  |  |  |
| *blaZ* A | 71 (16.6) | 111 (64.5) | <0.0001 | 3.881 | 3.059-4.935 |
| *blaZ* B | 70 (16.4) | 6 (3.5) | <0.0001 | 0.213 | 0.096-0.465 |
| *blaZ* C | 101 (23.7) | 43 (25) | 0.752 | 1.057 | 0.771-1.430 |
| *blaZ* D | 2 (0.5) | 0 | >0.999 | 0 | 0.000-4.732 |
| Unknown | 183 (42.9) | 12 (7) | <0.0001 | 0.163 | 0.093-0.279 |
| *agr*1 | 220 (51.5) | 43 (25) | <0.0001 | 0.485 | 0.366-0.632 |
| *agr*2 | 157 (36.8) | 23 (13.3) | <0.0001 | 0.364 | 0.242-0.536 |
| *agr*3 | 40 (9.4) | 105 (61) | <0.0001 | 6.517 | 4.757-8.966 |
| *agr*4 | 7 (1.6) | 0 | 0.201 | 0 | 0.000-1.343 |
| Unknown | 3 (0.7) | 1 (0.5) | >0.999 | 0.8275 | 0.119-5.720 |
| Univariate Association to *blaZ*, *agr* in PIE |  |  |  |  |  |
|  | PIE- | PIE+ | P-Value | RR |  |
| **Total N (%)** | 581 (97) | 18 (3) |  |  |  |
| *blaZ* A | 167 (28.7) | 15 (83.3) | <0.0001 | 2.899 | 2.075-3.515 |
| *blaZ* B | 76 (13.1) | 0 | 0.149 | 0 | 0.000-1.355 |
| *blaZ* C | 143 (24.6) | 1 (5.6) | 0.089 | 0.226 | 0.040-1.055 |
| *blaZ* D | 2 (0.3) | 0 | >0.999 | 0 | 0.000-57.77 |
| *blaZ* Unknown | 193 (33.2) | 2 (11.1) | 0.075 | 0.335 | 0.093-0.995 |
| *agr*1 | 260 (44.8) | 3 (16.7) | 0.027 | 0.372 | 0.130-0.882 |
| *agr*2 | 177 (30.5) | 3 (16.7) | 0.298 | 0.547 | 0.191-1.302 |
| *agr*3 | 133 (22.9) | 12 (66.7) | <0.0001 | 2.912 | 1.870-3.878 |
| *agr*4 | 7 (1.2) | 0 | >0.999 | 0 | 0.000-15.56 |
| *agr* Unknown | 4 (0.7) | 0 | >0.999 | 0 | 0.000-27.95 |

43.9% of isolates were *agr*1, 30.5% *agr*2, 24.2% *agr*3, 1.2% *agr*4, and 0.7% were unknown. *agr*1 and *agr*2 showed a relative increase from 2012 to 2019 by 12.3% and 26.1% respectively, while *agr*3 decrease by 31%. *agr*3 was significantly associated with HIE+ (p=<0.001, RR=6.517) and PIE+ (p=<0.001, RR=2.912). HIE+ and PIE+ were negatively associated with *agr1* but only HIE+ was negatively associated with *agr2* (Table 4).

A total of 9 unique CC lineages were identified with CC30 representing the plurality of isolates (21.5%). The proportion of CC30 isolates decreased across the study period (36.9% in 2012, 25.4% in 2013, 20.5% in 2014, and 22.4% in 2019). CC30 was associated with HIE+ (p=<0.001, RR=8.561) and PIE+ (p=0.003, RR=3.009). C45, CC5, CC8, and CC97 were negatively associated with HIE+.

There were multiple differences in the genetic makeup for HIE isolates with *blaZ* A and CC30 becoming significant predictors of HIE through logistic regression (Figure 5). Other *blaZ* types, *agr* types, and CCs were negative predictors of HIE on the basis that each isolate could only have one variant of *blaZ, agr, and* CC (e.g., absence of *agr3* was negatively associated with HIE). To further characterize how *blaZ*, *agr,* and CC30 interact, a logistic regression was performed with CC30, *blaZ* A, and *agr*3. These findings showed that CC30, *blaZ A,* and *agr*3 were strongly associated with each other suggesting co-localization. A combined category of *blaZ A* + *agr*3 was found to have the highest association with CC30 followed by the *agr*3 allele alone (Table 5). Given that only 1 isolate of CC30 did not have the *agr*3 allele, logistic regression could not be performed.

**Figure 5:**
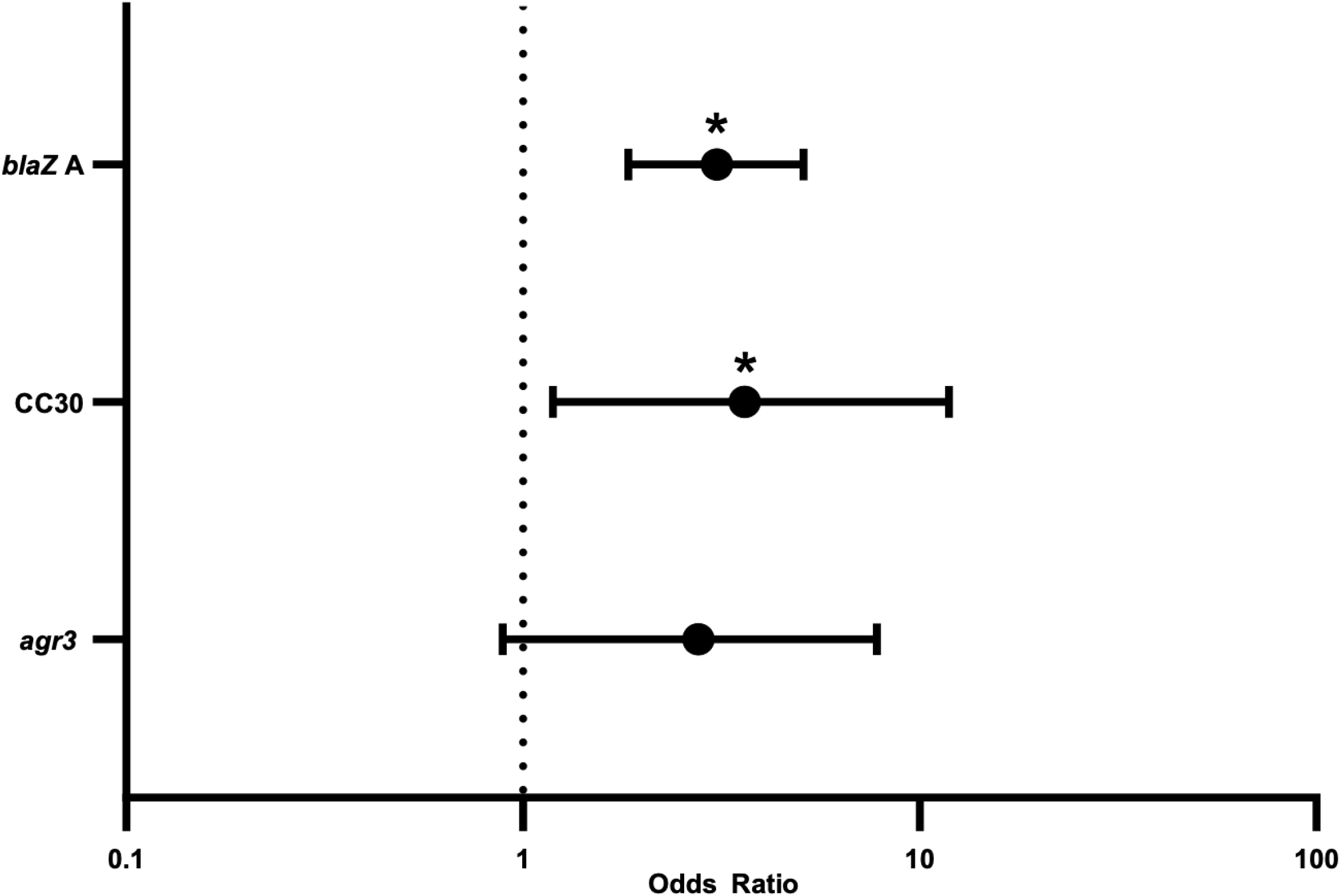
Genotypic features that are associated with the HIE through multivariate analysis.

**Table 5:**
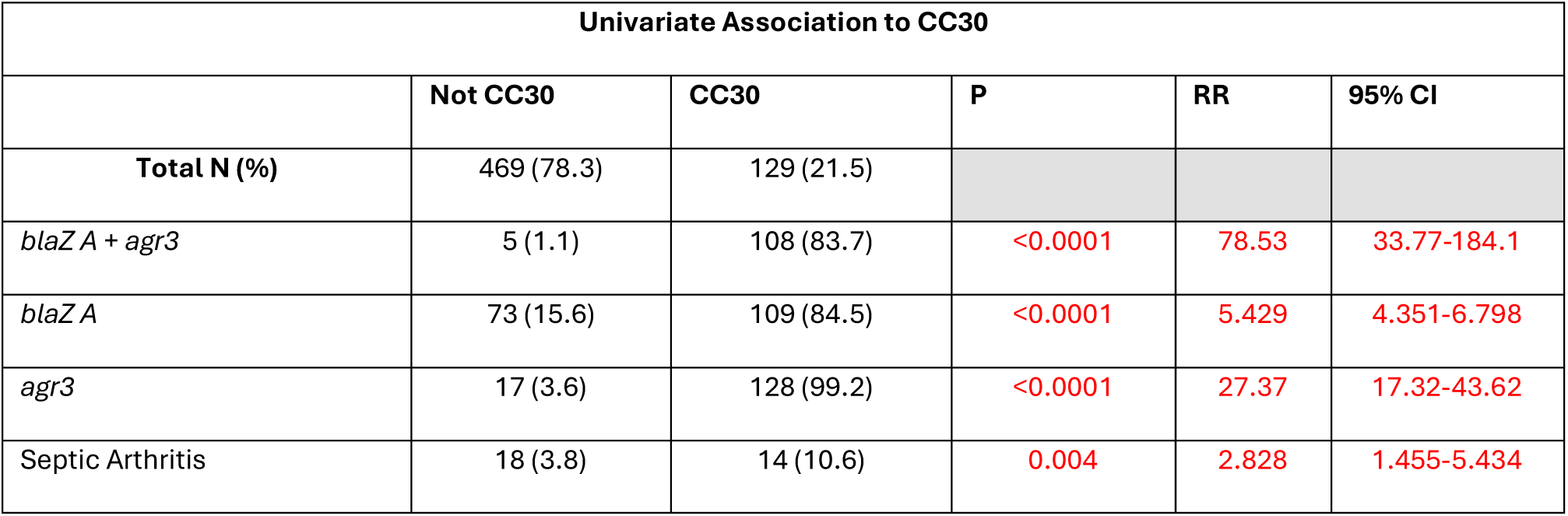
Features that are associated with CC30 through univariate analysis.

| Univariate Association to CC30 |  |  |  |  |  |
| --- | --- | --- | --- | --- | --- |
|  | Not CC30 | CC30 | P | RR | 95% CI |
| **Total N (%)** | 469 (78.3) | 129 (21.5) |  |  |  |
| *blaZ A + agr3* | 5 (1.1) | 108 (83.7) | <0.0001 | 78.53 | 33.77-184.1 |
| *blaZ A* | 73 (15.6) | 109 (84.5) | <0.0001 | 5.429 | 4.351-6.798 |
| *agr3* | 17 (3.6) | 128 (99.2) | <0.0001 | 27.37 | 17.32-43.62 |
| Septic Arthritis | 18 (3.8) | 14 (10.6) | 0.004 | 2.828 | 1.455-5.434 |

## Discussion

### Demographics of the HIE

The prevalence of HIE across studies varies based on geographic region and population. Utilizing the more common definition of a 4x increase in MIC at a higher level of inoculum (that used herein), the rates in our cohort appear to be similar to those observed in the United States (11,14), but substantially lower than South Korea (27) and South America (13). These later studies may be confounded by being limited to specific healthcare sites as opposed to entire regional population. The rates of HIE decreased across our study period from 38.9% in 2012 to 24.2% in 2019. Our rates of isolates with the PIE (12.2% of HIE+, 3.6% of all MSSA), were drastically lower than what has been previously reported in North America (PIE observed in 54.5% of isolates)(23).

Given possible adverse impacts of HIE on patient outcomes, we attempted to identify if patients infected by isolates with this phenotype could be predicted and if biomarkers for poor outcomes existed. Patient characteristics of those experiencing MSSA bacteremia were not different when comparing HIE+ versus HIE-, except for age and substance use. Additionally, such findings did not survive multivariable regression. These findings support no distinguishing characteristics that predispose to HIE status, similar to previous findings (23). HIE was not more commonly associated with either CA-, HCA-, and HA- acquisition. We infer this to mean that HIE cannot be reliably predicted from patient characteristics and that strains expressing the HIE phenotype *in-vitro* are not disproportionally associated with condition or exposure. Finally, we did not identify differentiating clinical biomarkers in those presenting with HIE+ MSSA BSI.

There was a significant increase in HIE phenotype in MSSA BSI isolates associated with “Septic Arthritis” infections. This remained significant after multivariate regression. Hematogenous complications are a known risk for CC30 MSSA (40) and isolates possessing virulence alleles such as *agr*3 (41). It may be that HIE is also associated with virulence factors that predispose to joint seeding.

### Outcomes and HIE

Our ability to assess how early therapy impacted bacteremia outcomes was limited owing to the myriad of antibiotic regimens patients received within 48hrs of presentation. Only once *S. aureus* is definitively identified, targeted anti-*Staphylococcal* therapy is instituted and only when susceptibilities confirm MSSA is vancomycin discontinued. The end effect is patients receiving multiple agents with anti-*Staphylococcal* activity for >24hrs.

We did not observe differences in either short- or long-term outcomes (i.e., time to clearance, recurrence risk, death) based on HIE status alone, or with specific targeted therapy (i.e., cefazolin vs cloxacillin). These findings contrast reports by Miller *et al.* (2018) and Lee *et al.* (2014), conducted in small cohorts with MSSA BSI from specific hospitals, demonstrating increased 30-day mortality and longer duration of BSI in HIE+. It is possible that exposure to multiple non-preferred agents with modest anti-*Staphylococcal* activity reduces the organism load in affected patients, abrogating the density dependent effect of partial beta-lactamase hydrolysis.

Some studies have observed different clinical outcomes based on HIE status, utilizing what we defined as PIE whose incidence in our cohort was so low that we were underpowered to detect a statistical difference (7,23). Kaplan-Meier curves (Figures 3&4) suggested a trend toward separation of the curves beyond 100-days, however this did not meet significance. Our survival analysis did not control for late palliation or refusing treatment, factors that could influence overall 1-year survival. We observed a non-significant trend towards increased risk of recurrence in HIE+ patients treated with cefazolin (inclusive of all recurrence episodes) as opposed to cloxacillin (where none were observed) (Supplemental Table 2). Despite the low prevalence of PIE in our cohort, we identified an association with increased 180-day mortality (p=0.024, RR=1.7, Supplemental Table 2) that approached the significance cutoff at 1-year (p=0.087, Supplemental Figure 1). This may suggest that clinical effects of HIE predominately manifest with PIE and that our population rates were too low to detect through HIE alone.

### Genomics and HIE

Univariate associations between HIE phenotypes showed positive association with *blaZ* A allele and negative associations with types B or unknown allele. This supports previously published data regarding *blaZ* A and HIE (7,11,27) but not *blaz* C, which was not significant in this study. There was a significant positive association between *agr* 3 and HIE but on regression *agr* 3 was no longer a significant predictor of HIE but was close to the predetermined significance cutoff (Figure 5, p=0.0598). *agr* 3 has been associated with HIE previously (15) and possible dysfunctional mutants (16), however, our genotypic analysis could not assess for *agr* function which may explain this difference.

Knowing that CC, *blaZ*, and *agr* alleles are not mutually exclusive we ran associations between these variables. We found that the strongest predictor of CC30 was the presence of *blaZ* A and *agr* 3 together (Table 5), suggesting that although these two alleles continue to independently associate with HIE, they are predominately found on MSSA belonging to CC30. It has been previously observed that CC30 are associated with these phenotypes (8,9) and by using WGS we were able to demonstrate that both alleles together associate with CC30.

Given the only clinical syndrome associated with HIE was septic arthritis, we also assessed if CC30 was associated with the same source. We found CC30 strains were associated with septic arthritis as a source (Table 5) with similar relative risk. CC30 has been linked to invasive infections in other studies (19,40,42,43) and this perhaps indicates an underlying virulence feature in HIE phenotypes that increases the odds of MSSA septic arthritis developing into bacteremia, or the reverse.

While the protocol (population-based, multi-center) and large population are clear strengths of this study, we acknowledge several limitations. For example, the inability to control confounding influences such as multiple antibiotic administration during initial presentation and WGS being restricted to 72% of the cohort’s isolate. It follows that reducing the inoculum through other agents may reduce the amount of beta-lactamase such that the amount of hydrolysis is negligible. Given this was not a randomized trial there is also the potential for treatment bias influencing which anti-*Staphylococcal* drug is given – such as patients with CNS complications requiring cloxacillin which was not coded in our data. Lastly, deaths were measured as all causes as opposed to attributable which could blur the influence of HIE.

## Conclusion

This represents the first non-selected, population-based study to assess the incidence, impact, and outcomes of HIE in persons experiencing MSSA BSI. The HIE decreased within the Calgary Health Zone from 36.9% to 22.4% across the study period. We did not find that the HIE phenotype can be predicted by patient characteristics of whom these isolates infect or clinical biomarkers (i.e., initial markers of disease severity). Neither short- nor long-term morbidity and mortality were significantly associated with the presence of the HIE, or the HIE in the context of targeted anti-Staphylococcal therapy unless specifically looking at PIE. It is possible that outcomes are influenced by early, empiric sub-optimal antimicrobials prior to targeted therapy with cefazolin.

## Data Availability

The data that support the findings of this study are available from the corresponding author, Dr. Michael D. Parkins, upon reasonable request.

## Funding

The study was supported by a grant from Alberta Precision Laboratories to M.D.P and D.B.G. The *Calgary BSI Cohort* is supported directly through funding from a Genome Canada 2017 Large scale applied research program award to I.L.

## Ethics

The study received ethics approval from the Conjoint Health Research Ethics Board of the University of Calgary (REB14-1456)

## Contributions

AJ Kipp=Methodology, Formal Analysis, Investigation, Visualization, Writing – Original Draft; K Du=Methodology, Formal Analysis, Writing – Review and Editing, B Waddell=Methodology, Formal Analysis, Writing – Review and Editing, S. Robinson= Methodology, Formal Analysis, Data curation, Writing – Review and Editing, J. Svishchuk= Methodology, Formal Analysis, Writing – Review and Editing; JM Conly=Project administration, Writing – Review and Editing; A. Ulke-Lemee= Methodology, Formal Analysis, Writing – Review and Editing; M Mapar= Methodology, Formal Analysis, Writing – Review and Editing; I. Lewis=Methodology, Formal analysis, Resources, Funding Acquisition; DB Gregson=Conceptualization, Resources, Data Curation, Writing – Review and Editing, Project Administration, Writing – Review and Editing, Funding Acquisition; MD Parkins= Conceptualization, Resources, Writing – Review and Editing, Supervision, Project Administration, Funding Acquisition

## Notes

### Competing Interest Statement

The authors have declared no competing interest.

### Funding Statement

This study was supported by funding from Alberta Precision Laboratories and Genome Alberta

### Author Declarations

Approval for this study was granted by University of Calgary's Conjoint Regional Health Ethics Board.

